# Effects of participatory organizational interventions on mental health and work performance: A protocol for systematic review and meta-analysis

**DOI:** 10.1101/2023.06.16.23290695

**Authors:** Mako Iida, Asuka Sakuraya, Kotaro Imamura, Hiroki Asaoka, Hideaki Arima, Emiko Ando, Akiomi Inoue, Reiko Inoue, Mai Iwanaga, Hisashi Eguchi, Yasumasa Otsuka, Yuka Kobayashi, Yu Komase, Kazuto Kuribayashi, Natsu Sasaki, Kanami Tsuno, Ayako Hino, Kazuhiro Watanabe, Takeshi Ebara, Akihito Shimazu, Norito Kawakami, Akizumi Tsutsumi

## Abstract

**Introduction:** Participatory organizational interventions to improve psychosocial working conditions are important for a safe and healthy work environment. However, there are few systematic reviews or meta-analyses investigating the effects of these interventions on workers’ mental health and work-related outcomes. We apply the protocol for systematic review and meta-analysis to examine the effect of participatory organizational intervention on mental health and work performance.

**Methods and analysis:** The participants, interventions, comparisons, and outcomes (PICO) of the studies in this systematic review and meta-analysis were defined as follows: (P) inclusion of all workers, (I) participatory organizational intervention, (C) treatment as usual or no intervention (including waitlist control), and (O) mental health and work performance. Published studies will be searched using the following electronic databases: PubMed, EMBASE, PsycINFO, PsycARTICLES, and Japan Medical Abstracts Society. Study selection and the risk of bias assessment will be performed independently by two reviewers. A meta-analysis will be performed to statistically synthesize the included studies.

**Ethics and dissemination:** This study does not require ethical approval. The findings and results will be submitted to and published in a scientific peer-reviewed journal.

**Registration details:** The study protocol was registered at the UMIN registry (registration number: UMIN000049453).

## Introduction

Negative work conditions such as a hazardous work environment and work organization, harmful or poor work conditions, or poor work relationships can significantly deteriorate the well-being of workers (1). To promote and maintain the highest degree of physical, mental, and social well-being of workers in all occupations, it is essential to guarantee a secure and healthy workplace for all workers (2-4). For a safe and healthy work environment, it is important to have organizational interventions to improve psychosocial working conditions (1, 5-8). Organizational interventions are “planned actions that primarily directly target working conditions with the aim of promoting and maintaining the highest degree of physical, mental, and social well-being of workers in all occupations” (9).

Numerous scholars have highly suggested involving workers in implementing organizational interventions (8, 10-13). For instance, the International Organization for Standardization (ISO) 45003 guideline suggests that the participation of workers is essential for developing, planning, implementing, maintaining, evaluating, and continually improving healthy and safe workplaces and managing psychosocial risk (13). Since worker participation increases their control, sense of fairness, justice, and support; their involvement also helps optimize the intervention’s fit to the organizational culture and context, and thus, promoting worker participation is a necessary component of organizational intervention (8). Sakuraya et al. defined participatory organizational intervention as “workers participate on steps of an intervention, such as action planning, implementing, evaluating, and reviewing the intervention” (9).

Several cluster randomized controlled trials (cRCTs) revealed the effect of participatory organizational interventions. For mental health outcomes, the participatory organizational intervention has effects on improved minor psychiatric morbidity (General Health Questionnaire) (14) and depersonalization (subscale of burnout) (15). For work-related outcomes, psychosocial work environment (e.g., coworker support and well-defined and realistic workplace goal) (16), sickness absence (17), and job performance (14) were also improved by participatory organizational intervention. Thus, participatory organizational intervention may be promising for enhancing mental health and work-related outcomes.

There are few systematic reviews or meta-analyses that gather evidence from cRCTs on the effects of participatory organizational intervention on workers’ mental health and work-related outcomes. In previous systematic reviews, organizational interventions had favorable impacts on mental health (e.g., decreased distress or burnout) (5, 7) and work-related outcomes (e.g., absenteeism, sickness absence) (5, 6). However, these reviews included non-participatory organizational interventions as well as participatory ones. Therefore, the effectiveness of the participatory organizational intervention itself is unclear. Also, one meta-analysis reported a non-significant effect of participatory organizational interventions on stress level (e.g., occupational stress or burnout) (standardized mean difference (SMD) -0.12; 95% CI -0.30 to 0.05) (18). However, this study targeted only healthcare workers, thus, the effects of participatory organizational intervention among general workers are unknown.

Therefore, the objective of this study is to examine the effect of participatory organizational intervention on mental health and work performance among all workers. This review treats outcomes related to workers’ well-being or productivity. To the best of our knowledge, this will be the first systematic review and meta-analysis conducted specifically to investigate the effect of participatory organizational interventions among workers.

## Methods and analysis

### Study design

This is a systematic review and meta-analysis protocol for cRCT studies, following the Preferred Reporting Items for Systematic Reviews and Meta-Analysis protocols (PRISMA-P) guideline (19) (Appendix 1). The systematic review and meta-analysis will be reported according to the PRISMA (Preferred Reporting Items for Systematic Reviews and Meta-Analysis) 2020 guideline (20). The study protocol was registered at the UMIN registry (registration number: UMIN000049453).

### Eligibility criteria

The participants, interventions, comparisons, and outcomes (PICO) of the studies in this systematic review and meta-analysis were defined as follows: (P) inclusion of all workers, (I) participatory organizational intervention, (C) treatment as usual or no intervention (including waitlist control), and (O) mental health and work performance. The definition of participatory organizational intervention was followed by an opinion paper by Sakuraya et al. (2023) (9). For mental health outcomes, positive mental health (e.g., optimism, satisfaction, positive affect, well-being, or work engagement), or mental health conditions (e.g., mental disorders, depression, burnout, or stress) will be included. In addition, work performance outcomes, such as work capacity evaluation, effectiveness, or presenteeism, will be included. There will be no exclusion criteria for workers based on their employment status, job type, and shift type.

The inclusion criteria are as follows:

1. Studies that included participatory organizational intervention
2. Studies that included participants who were working as of the baseline survey period
3. Studies that assessed mental health or work performance outcomes
4. Studies that used a cRCT design
5. Studies published in English or Japanese
6. Studies published in peer-reviewed journals (including advanced online publication).

Information source, search strategy, and data management Published studies will be searched using the following electronic databases: PubMed, EMBASE, PsycINFO, PsycARTICLES, and Japan Medical Abstracts Society. The search terms will include words related to the PICO of the studies. The search strategy is shown in Appendix 2. All identified studies will be managed within both EndNote-20 Library and Microsoft Excel (Washington, USA) files. Before screening the studies, MIi will remove duplicate entries by using EndNote-20 Library and export the data to the Excel file.

### Study selection process

We plan to outsource some of the sifting work to specialist contractors. A total of 15 investigators (MIi, ASa, KI, HAs, EA, AI, RI, MIw, HE, YO, YKob, YKom, NS, KT, KW), and a specialist contractor representative will independently conduct the screening of studies according to the eligibility criteria created earlier in the shifting phase, and the full text of all eligible studies will be obtained. In the full-text review phase, the full texts will be reviewed using a standardized form for assessing eligibility for this study. When resolution cannot be accomplished, the disagreements will be settled by consensus with discussion among all authors. Corresponding authors will be contacted directly if (1) the publication is unclear and may be related to multiple interpretations, or (2) the collected data from the publication did not show data relevant to our study analyses. The reasons for excluding studies will be recorded. A flow chart will be provided to show the entire review process.

### Data collection

Data will be extracted independently from the included studies by 15 investigators (MIi, ASa, KI, HAs, EA, AI, RI, MIw, HE, YO, YKob, YKom, NS, KT, KW) using a standardized data extraction form. All authors will consult and reach an agreement to resolve any disagreements or inconsistencies. Data of the year of publication, the country in which the study was conducted, the length of follow-up, sample size, demographic characteristics of participants, the contents of intervention, condition of the control group, and outcome variables, and result on mental health or work performance outcomes will be extracted. This extraction form will be piloted and adjusted as needed. Means and standard deviations (SDs) of outcomes at baseline and post-intervention surveys, as well as the number of participants at analyses of intervention and control groups, will be collected for the meta-analysis.

### Risk of bias in individual studies and assessment of metabias

A total of 15 investigators (MIi, ASa, KI, HAs,EA, AI, RI, MIw, HE, YO, YKob, YKom, NS, KT, KW) will independently assess the study quality of each selected study using the risk of bias assessment tool of the Grading of Recommendation Assessment, Development, and Evaluation (GRADE) system (21, 22). The risk of bias assessment tool of the GRADE system evaluates a cluster-randomized controlled study based on six domains: 1a) bias arising from the randomization process; 1b) bias arising from the\ timing of identification or recruitment of participants, 2) bias due to deviations from intended interventions; 3) bias due to missing outcome data; 4) bias in measurement of the outcome; and 5) bias in selection of the reported result. Each item will be assessed as a low risk of bias; some concerns; or a high risk of bias. Disputes between the 15evaluators will be resolved by all authors until a consensus is reached. A summary of findings will be created using the GRADE approach to grade the certainty of evidence. Publication bias will be assessed for meta-bias using Egger’s test as well as visually on a funnel plot.

## Data synthesis and statistical methods

### Primary analyses

For the main analyses, the included studies will be statistically synthesized by a meta-analysis to estimate the pooled effects of the participatory organizational interventions on mental health and work performance outcomes, respectively. The continuous outcomes will be synthesized by calculating SMDs and their 95% confidence intervals. If the included studies use dichotomous and continuous variables, the continuous outcomes will be converted to dichotomous variables based on appropriate cutoff points and synthesized by calculating odds ratios (ORs) or relative risks (RRs) and their 95% confidence intervals. If there is no reasonable cutoff point, we will analyze dichotomous variables and continuous variables separately. If a meta-analysis cannot be conducted because only two or fewer studies are eligible and included, the findings will be presented in narrative form. If no heterogeneity is observed (e.g., types of interventions or populations), a fixed-effect model will be used, and otherwise, a random-effects model will be used. We will assess the heterogeneity by using the Q statistic (23).

All the collected data and analyzed results will be deposited by the corresponding author and available upon requests by external reviewers.

### Subgroup and sensitivity analyses

Subgroup analyses will be conducted to compare the results under specific contents of intervention (e.g., All workers participate in every step of the intervention vs. not) and outcome (e.g., positive mental health/mental health conditions), if enough data to conduct such analyses can be collected. Any differences between subgroups will be reported, and our findings will be explained in light of these differences. For included studies with a GRADE of low risk, a sensitivity analysis will be conducted (21).

### Patient and public involvement

This study has had no direct patient or public involvement in its design.

### Ethics and dissemination

Because this systematic review and meta-analysis is based on previously published studies, it does not require ethical approval. The findings and results will be submitted to and published in a scientific peer-reviewed journal.

## Strengths and limitations

To our knowledge, this will be the first systematic review and meta-analysis to reveal integrated evidence for the effect of participatory organizational intervention on workers’ mental health and work performance. This research will demonstrate how this type of intervention can impact the well-being and productivity of workers. Considering the importance of well-being and productivity at a workplace, the findings of this study will be useful for public and occupational health.

However, this systematic review and meta-analysis study may have some limitations. The generalization of the findings may be restricted based on the demographic characteristics of the participants included in the selected studies. Furthermore, the selection of databases for this review is adopted on the basis of previous studies, and we are not able to make a comprehensive search. For example, articles in languages other than English or Japanese, and gray literature, such as conference proceedings and unpublished manuscripts, are not included in this review.

## Data Availability

No data included in the present study because this is a protocol for systematic review and meta-analysis.

## Authors’ contributions

MIi, ASa, KI, HAs, HAr, EA, AI, RI, MIw, HE, YO, YKob, YKom, KK, NS, KT, AH, KW, ASh, NK, and AT have made substantial contributions to the conception and design. ASh, NK, and AT supervised the research plan. MIi and ASa prepared the first draft. KI, HAs, HAr, EA, AI, RI, MIw, HE, YO, YKob, YKom, KK, NS, KT, AH, KW, TE, ASh, NK, and AT revised it critically for important intellectual content, and all authors approved the final version to be published. KI, HE, and TE obtained funding for the study.

