## Appendix 2 for "Effects of participatory organizational interventions on mental health and work performance: A protocol for systematic review and meta-analysis"

**Supplementary Appendix 2**

Search terms for PubMed

("Occupational Groups"[Mesh] OR "Occupational Health"[Mesh] OR “enterprise*”[tiab] OR "business*"[tiab] OR "employed"[tiab] OR "employee*"[tiab] OR "employer*"[tiab] OR "employment*"[tiab] OR "informal sector*"[tiab] OR "informal work*"[tiab] OR "laborer*"[tiab] OR "labourer*"[tiab] OR "occupation*"[tiab] OR "personnel*"[tiab] OR "professional*"[tiab] OR "staff*"[tiab] OR "vocation*"[tiab] OR "worker*"[tiab] OR "workforce"[tiab] OR "workplace*"[tiab] OR "work place*"[tiab] OR "worksite*"[tiab] OR "work site*"[tiab] OR "aide*"[tiab] OR "audiologist*"[tiab] OR "ambulance*"[tiab] OR "care assistant*"[tiab] OR "clinician*"[tiab] OR "coastguard*"[tiab] OR "coast guard*"[tiab] OR "dentist*"[tiab] OR "detective*"[tiab] OR "disaster responder*"[tiab] OR "doctor*"[tiab] OR "domestic health car*"[tiab] OR "domiciliary car*"[tiab] OR "emergency service*"[tiab] OR "emergency responder*"[tiab] OR "emergency medical"[tiab] OR "firefighter*"[tiab] OR "fire fighter*"[tiab] OR "first responder*"[tiab] OR "general practitioner*"[tiab] OR "health care provider*"[tiab] OR "healthcare provider*"[tiab] OR "health visitor*"[tiab] OR "home care service*"[tiab] OR "humanitarian aid*"[tiab] OR "humanitarian relie*"[tiab] OR "humanitarian service*"[tiab] OR "law enforc*"[tiab] OR "lifeguard*"[tiab] OR "life guard*"[tiab] OR "medical resident*"[tiab] OR "medic"[tiab] OR "medics"[tiab] OR "nurse*"[tiab] OR "nursing"[tiab] OR "midwi*"[tiab] OR "paramedic*"[tiab] OR "policemen"[tiab] OR "policeman"[tiab] OR "police men"[tiab] OR "police man"[tiab] OR "police women"[tiab] OR "police woman"[tiab] OR "police officer*"[tiab] OR "firemen"[tiab] OR "fireman"[tiab] OR "fire men"[tiab] OR "fire man"[tiab] OR "fire women"[tiab] OR "fire woman"[tiab] OR "pharmacis*"[tiab] OR "psychologist*"[tiab] OR "physician*"[tiab] OR "practitioner*"[tiab] OR "relief work*"[tiab] OR "rescuer*"[tiab] OR "rescue work*"[tiab] OR "therapist*"[tiab] OR "veterinaria*"[tiab])

AND

("Employee Grievances"[Mesh] OR "Work Schedule Tolerance"[Mesh]OR "Bullying"[Mesh] OR "Interpersonal Relations"[Mesh] OR "Prejudice"[Mesh] OR "Social Discrimination"[Mesh] OR "harassment, non-sexual"[Mesh] OR “risk management”[Mesh] OR "Organizational Culture"[Mesh] OR “Organizational Policy”[MeSH] OR “leadership”[Mesh] OR "Personnel Downsizing"[Mesh] OR "Staff Development"[Mesh] OR "Employee Performance Appraisal"[Mesh] OR “inservice training*”[tiab] OR “in service training*”[tiab] OR "organizational intervention*"[tiab] OR "organisational intervention*"[tiab] OR “participatory intervention*”[tiab] OR “team intervention*”[tiab] OR “compressed hour*”[tiab] OR "compressed work*"[tiab] OR “compressed week*”[tiab] OR “day-time”[Tiab] OR “daytime”[tiab] OR “day time”[tiab] OR “flexible schedule*”[tiab] OR “flexible work*”[tiab] OR “inflexible schedule*”[tiab] OR “inflexible work*”[tiab] OR “full-time”[Tiab] OR “gig economy”[tiab] OR “long hour*”[tiab] OR “night-time”[Tiab] OR “nighttime”[tiab] OR “night time”[tiab] OR “night shift*”[tiab] OR “overtime*”[tiab] OR “part-time”[Tiab] OR “recovery”[tiab] OR “remote work*”[tiab] OR “shift work*”[Tiab] OR “self-scheduling”[tiab] OR “temporary work*”[Tiab] OR “work schedule*”[tiab] OR “working schedule*”[tiab] OR “working hour*”[Tiab] OR “work hour*” [tiab] OR “working time”[Tiab] OR “work shift*”[Tiab] OR “zero hour*”[tiab] OR ("life"[tiab] AND “balance"[tiab]) OR (“work*”[tiab] AND "life"[tiab] AND “balance"[tiab]) OR ("life"[tiab] AND “family"[tiab]) OR ("reconciling"[tiab] AND “work*"[tiab]) OR ("living"[tiab] AND "working"[tiab]) OR “work overload*”[Tiab] OR “work over-load*”[Tiab] OR “work pace”[tiab] OR “time pressure*”[Tiab] OR “decision latitude”[Tiab] OR “demand resource*”[Tiab ] OR “effort reward*”[Tiab] OR “high demand*”[Tiab] OR “job control”[Tiab] OR “job demand*”[Tiab] OR “job strain”[Tiab] OR “lack of control”[Tiab] OR “task restructur*”[tiab] OR “low control”[Tiab] OR “work demand*”[Tiab] OR “work control”[Tiab] OR “work influence*”[Tiab] OR “work strain”[Tiab] OR “boredom”[Tiab] OR “coping”[Tiab] OR ((“control”[tiab] OR “unpleasant”[tiab] OR “aversive”[tiab]) AND “task*”[tiab]) OR “job content”[tiab] OR monoton*[tiab] OR “under stimulat*”[tiab] OR “ageism”[Tiab] OR “aggression”[tiab] OR “bullying”[Tiab] OR “discrimination”[Tiab] OR “interpersonal relation*”[Tiab] OR “harass*”[Tiab] OR “homophobia”[Tiab] OR “microaggression”[tiab] OR “prejudice”[tiab] OR “racism”[Tiab] OR “sexism”[Tiab] OR “silent workplace*”[Tiab] OR “social capital” [Tiab] OR “solitary work*” [tiab] OR “isolated work*” [tiab] OR “supervision”[tiab] “victimization*”[Tiab] OR “work place conflict*”[Tiab] OR “workplace violen*”[Tiab] OR “work place violen*”[Tiab] OR "lean management"[tiab] OR "risk management"[tiab] OR “safety management”[tiab] OR “work environment”[tiab] OR “working environment”[tiab] OR “work condition*”[tiab] OR “working condition*”[tiab] OR “work organization”[tiab] OR “work design”[tiab] OR “communication*”[tiab] OR “organisational culture”[tiab] OR “organizational culture”[tiab] OR “organisational function”[tiab] OR “organizational function”[tiab] OR “organisational injustice*”[Tiab] OR “organizational injustice*”[Tiab] OR “health and safety”[tiab] OR “organisational justice*”[Tiab] OR “organizational justice*”[Tiab] OR “leadership”[tiab] OR “lean production”[Tiab] OR "lean management"[tiab] OR “labour relation*”[tiab] OR “management practice*”[tiab] OR “management culture”[tiab] OR “management measure*”[tiab] OR ((“participation”[tiab] OR “involvement”[tiab]) AND “decision making*”[tiab]) OR “organisational policy”[tiab] OR “organizational policy”[tiab] OR “organisational policies”[tiab] OR “organizational policies”[tiab] OR “psychosocial risk*”[tiab] OR “procedural justice”[tiab] OR “procedural injustice”[tiab] OR “clear role”[tiab] OR “skill discretion*”[Tiab] OR “role ambiguity”[Tiab] OR “role conflict*”[Tiab] OR “role clarity”[tiab] OR “unclear role”[tiab] OR “work role*”[Tiab] OR “career development”[tiab] OR “job security”[Tiab] OR “job insecurity”[Tiab] OR “over skilled”[tiab] OR “performance evaluation”[tiab] OR “professional development”[tiab] OR ((“work”[tiab] OR “job”[tiab]) AND “promotion”[tiab]) OR “staff development”[Tiab] OR “under skilled”[tiab] OR “team building”[tiab] OR “teambuilding”[tiab] OR (“team*”[tiab] AND “participatory”[tiab]) OR “teamwork*”[tiab] OR ((“team*”[tiab] OR “co worker”[tiab] OR “colleague*”[tiab] ) AND (“work”[tiab] OR “building”[tiab] OR “program*”[tiab])))

AND

(randomized controlled trial [pt] OR (randomized [tiab] AND controlled [tiab] AND trial [tiab]))

AND

(("Mental Disorders"[Mesh] OR "Mental Health"[Mesh] OR "Psychology, Industrial"[Mesh] OR "Stress, Psychological"[Mesh] OR "adjustment"[tiab] OR "affective disorder*"[tiab] OR "anxiet*"[tiab] OR "bipolar*"[tiab] OR "burn out*"[tiab] OR "burnout*"[tiab] OR "CMD" [tiab] OR "depressi*"[tiab] OR "eating disorder*"[tiab] OR "mental disorder*"[tiab] OR "mental health*"[tiab] OR "mental illness*"[tiab] OR "mood disorder*"[tiab] OR "obsessive compulsive disorder*"[tiab] OR "ocd"[tiab] OR "panic disorder*"[tiab] OR "phobi*"[tiab] OR "post traumatic*"[tiab] OR "psychiatric diagnos*"[tiab] OR "psychiatric disease*"[tiab] OR "psychiatric disorder*"[tiab] OR "psychiatric illness*"[tiab] OR "psychological disorder*"[tiab] OR "psychos*"[tiab] OR "psychotic*"[tiab] OR "psychological distress*"[tiab] OR "ptsd"[tiab] OR "ptss"[tiab] OR "somatoform disorder*"[tiab] OR "schizophren*"[tiab] OR "stress*"[tiab]) OR

("Optimism"[Mesh] OR "Personal Satisfaction"[Mesh] OR "Self Concept"[Mesh:NoExp] OR "Self Efficacy"[Mesh] OR "Self-Control"[Mesh] OR "life engag*"[tiab] OR "life satisf*"[tiab] OR "meaning of life"[tiab] OR "purpose in life"[tiab] OR "positive affect*"[tiab] OR "positive emotion*"[tiab] OR "resilien*"[tiab] OR "self concept*"[tiab] OR "self control*"[tiab] OR "self efficac*"[tiab] OR "self esteem*"[tiab] OR "swb"[tiab] OR "well being*"[tiab] OR "wellbeing*"[tiab] 　OR "Job Satisfaction"[MeSH] OR "job satisf*"[tiab] OR "work satisf*"[tiab] OR "work engag*"[tiab]) OR

("Work Capacity Evaluation"[Mesh] OR "effectiveness"[tiab] OR "employabil*"[tiab] OR "presenteeism*"[tiab] OR "productivit*"[tiab] OR "work abilit*"[tiab] OR "work capacit*"[tiab] OR "work disabilit*"[tiab] OR "work function*"[tiab] OR "work participati*"[tiab] OR "work performan*"[tiab]))
